# Use of Artificial Intelligence on spatio-temporal data to generate insights during COVID-19 pandemic: A Review

**DOI:** 10.1101/2020.11.22.20232959

**Authors:** Gihan Jayatilaka, Jameel Hassan, Umar Marikkar, Rumali Perera, Suren Sritharan, Harshana Weligampola, Mevan Ekanayake, Roshan Godaliyadda, Parakrama Ekanayake, Vijitha Herath, G M Dilshan Godaliyadda, Anuruddhika Rathnayake, Samath D. Dharmaratne, Janaka Ekanayake

## Abstract

The COVID-19 pandemic, within a short time span, has had a significant impact on every aspect of life in almost every country on the planet. As it evolved from a local epidemic isolated to certain regions of China, to the deadliest pandemic since the influenza outbreak of 1918, scientists all over the world have only amplified their efforts to combat it. In that battle, Artificial Intelligence, or AI, with its wide ranging capabilities and versatility, has played a vital role and thus has had a sizable impact. In this review, we present a comprehensive analysis of the use of AI techniques for spatio-temporal modeling and forecasting and impact modeling on diverse populations as it relates to COVID-19. Furthermore, we catalogue the articles in these areas based on spatio-temporal modeling, intrinsic parameters, extrinsic parameters, dynamic parameters and multivariate inputs (to ascertain the penetration of AI usage in each sub area). The manner in which AI is used and the associated techniques utilized vary for each body of work. Majority of articles use deep learning models, compartment models, stochastic methods and numerous statistical methods. We conclude by listing potential paths of research for which AI based techniques can be used for greater impact in tackling the pandemic.

## 1. Introduction

The COVID-19 pandemic caused by the novel coronavirus (SARS-CoV-2) exemplifies the vulnerabilities of healthcare systems in the face of unforeseen adversity. After COVID-19 emerged from China in January 2020, it has spread across the globe and has been responsible for the ongoing pandemic that has claimed hundreds of thousands of lives around the world (as of September 2021, 4.5 million deaths have been reported^1^). Artificial Intelligence (AI) is at the forefront in the global response to the prevailing COVID-19 pandemic. In particular, AI solutions have been developed for infection detection, forecasting, response planning, recovery planning, risk assessment and patient prioritization, screening and diagnosis, contact tracing, understanding social interventions, and automated patient care.^2, 3, 4^

AI is the notion of mimicking human intellect on a computer, through a collection of operations. The concept of AI has been with us since the early 1900s and was formalized by Alan Turing in the 1950s. However, up until the 21st century AI was at a nascent state primarily due to the limited availability of processing power. The AI revolution as we know came about in the early 2010s when AlexNet,^5^ an artificial deep neural network-based solution, managed to triumph the competition in an image classification challenge. Since then, AI has permeated almost every scientific discipline including agriculture, education, manufacturing, finance, transportation, media, and healthcare. Global Health is another key sector which is widely influenced by AI. In fact, AI systems assist healthcare by performing tasks such as diagnosis, risk assessment, forecasting and surveillance, and health policy and planning.^6, 7^

The furtherance of AI systems has been supplemented by the abundance of data, i.e. Big Data, that is available to researchers in the era of COVID-19. Hence, hundreds of articles on AI-based systems for different tasks in the fight against COVID-19 are published constantly (See Fig. 1). To assist further research in the field, this plethora of articles have been summarized and evaluated in several review articles.^2, 6, 8, 9, 10, 11, 12^ These review articles discuss the impact of AI in a general sense i.e. without further categorization into specific topics. However, with further proliferation of articles, a need for more rigorous review has emerged, specifically, reviews that catalogue projects based on their specific utilization of AI i.e. cataloguing based on which tasks are achieved by AI and how. In this paper, we identify and categorize papers into two primary categories based on the tasks achieved through AI -- 1) spatio-temporal modeling and forecasting where features like geographical location and time are utilized for modeling and forecasting, 2) modeling the impact on diverse populations where additional features like age, medical conditions, environmental differences, socio-economic dependence, political dependence, population flow, and travel patterns are utilized for modeling. In this review, we avert the normative style of review articles on AI systems for COVID-19 and present an in-depth analysis focusing on the aforementioned two tasks. We also discuss the potential of AI systems and how the worldwide AI community should respond to COVID-19 in the near term as well as how we can learn from our experiences for future pandemics.

**Figure 1:**
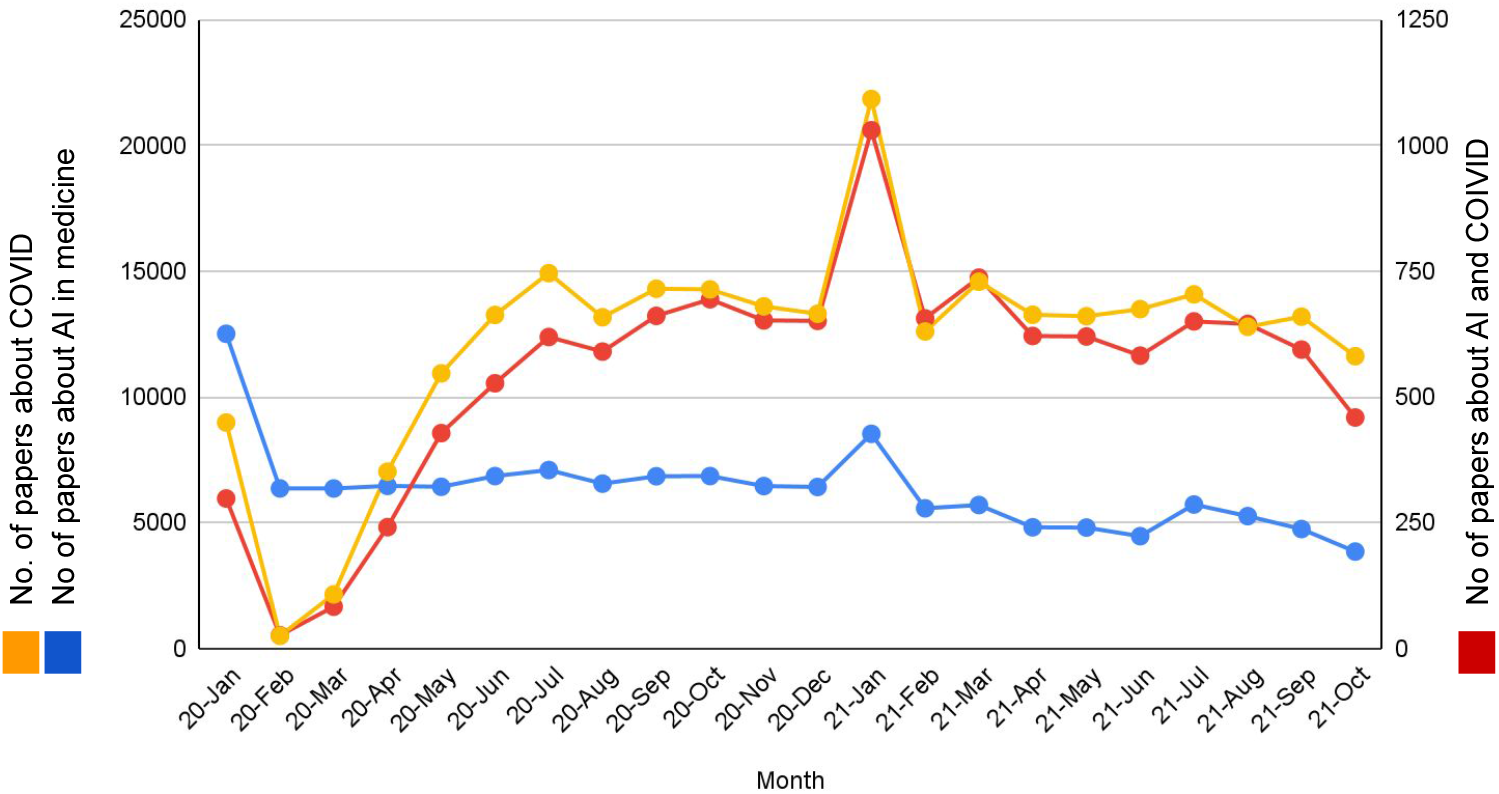
Time variation of number of papers in AI (for medical applications only), COVID and AI+COVID

## 2. Search Strategy and Selection Criteria

The literature survey for research papers on existing work on COVID-19 (or pandemics in general) and AI was done using IEEE Xplore, PubMed and Google Scholar for articles published between the time period 2019 and 2021 September. Only articles written in English were considered in this review. The keywords used were [”COVID-19”, “pandemic” or “sars-cov-2”] and [”AI” or “artificial intelligence” or “machine learning” or “pattern recognition” or “prediction” or “forecasting”] and [”time” or “location”, “demography” or “country” or “spatio-temporal”]. Papers were shortlisted from peer reviewed papers published or in press for journals indexed in Scimago. These criteria resulted in 156 research papers. The texts were examined to pick 68 research papers that matched the scope of this survey. This is illustrated in Fig. 2.

**Figure 2:**
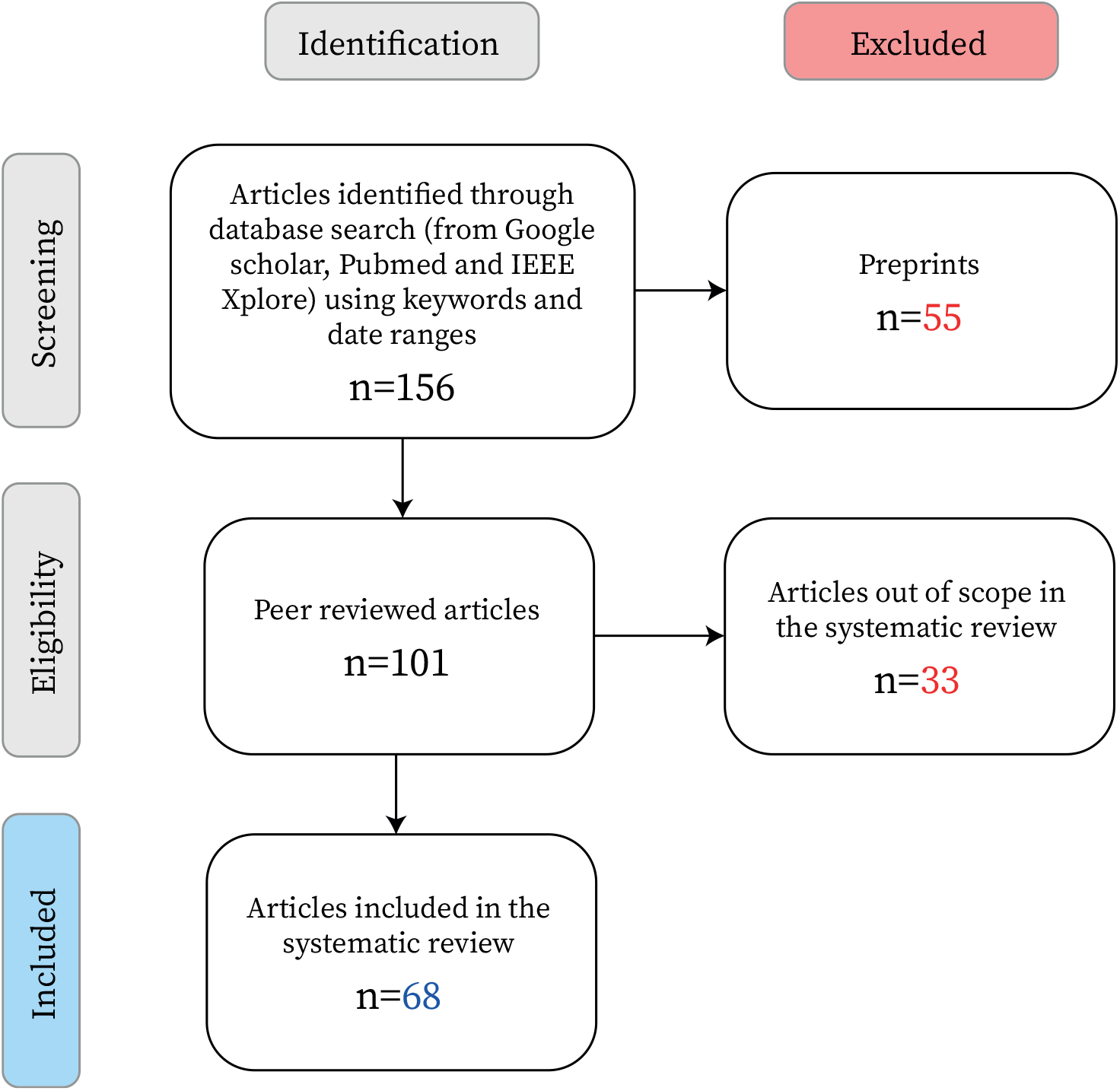
Flowchart for the literature survey methodology

## 3. Results

Our literature survey showed that spatio-temporal forecasting was performed either as a time series forecasting problem or as a diversity aware analysis by taking the diversity of populations into account via one/multiple parameter(s). The parameters being used are split into groups based on whether the parameter is inherent to the individuals (intrinsic), the environment (extrinsic) or an activity performed by the individuals (dynamic).The papers are grouped into sections depending on the parameters used to model the affected parties as given in Table 1.

**Table 1:** Parameters being used to characterize the diversity in population in the literature

| Category | Parameter | No. of papers | Section |
| --- | --- | --- | --- |
|  | Unparameterized | 22 | Section 3.1 |
| Intrinsic | Medical conditions | 2 | Section 3.2.1 |
|  | Age | 3 |  |
| Extrinsic | Environmental | 4 | Section 3.2.2 |
|  | Socio-Economic-Political | 8 |  |
| Dynamic | Traveling and Population flow | 5 | Section 3.2.3 |
| Multi | Multiple-parameters | 5 | Section 3.2.4 |
|  | **Total** | **49** |  |

### 3.1. Spatio-Temporal modeling and forecasting

The first cluster of research papers on modeling and forecasting was based on spatial and temporal data alone. This enabled us to identify the impact of AI usage on spatio-temporal data to facilitate the fight against COVID-19 by obtaining predictions on the number of total patients in the future, thus giving insight on the trajectory a country/state is heading in. It facilitates the future allocation of hospital resources, the lack of which has caused widespread commotion in many countries.^13, 14^ This data is also crucial in instructing the authorities involved in policy and decision making, and will thereby ensure efficient containment of the virus spread.^15^ Several methods including statistical approaches, machine learning techniques, deep learning models, and time series models have been evaluated in this landscape. The various AI models used and example papers are listed in Fig. 3.

**Figure 3:**
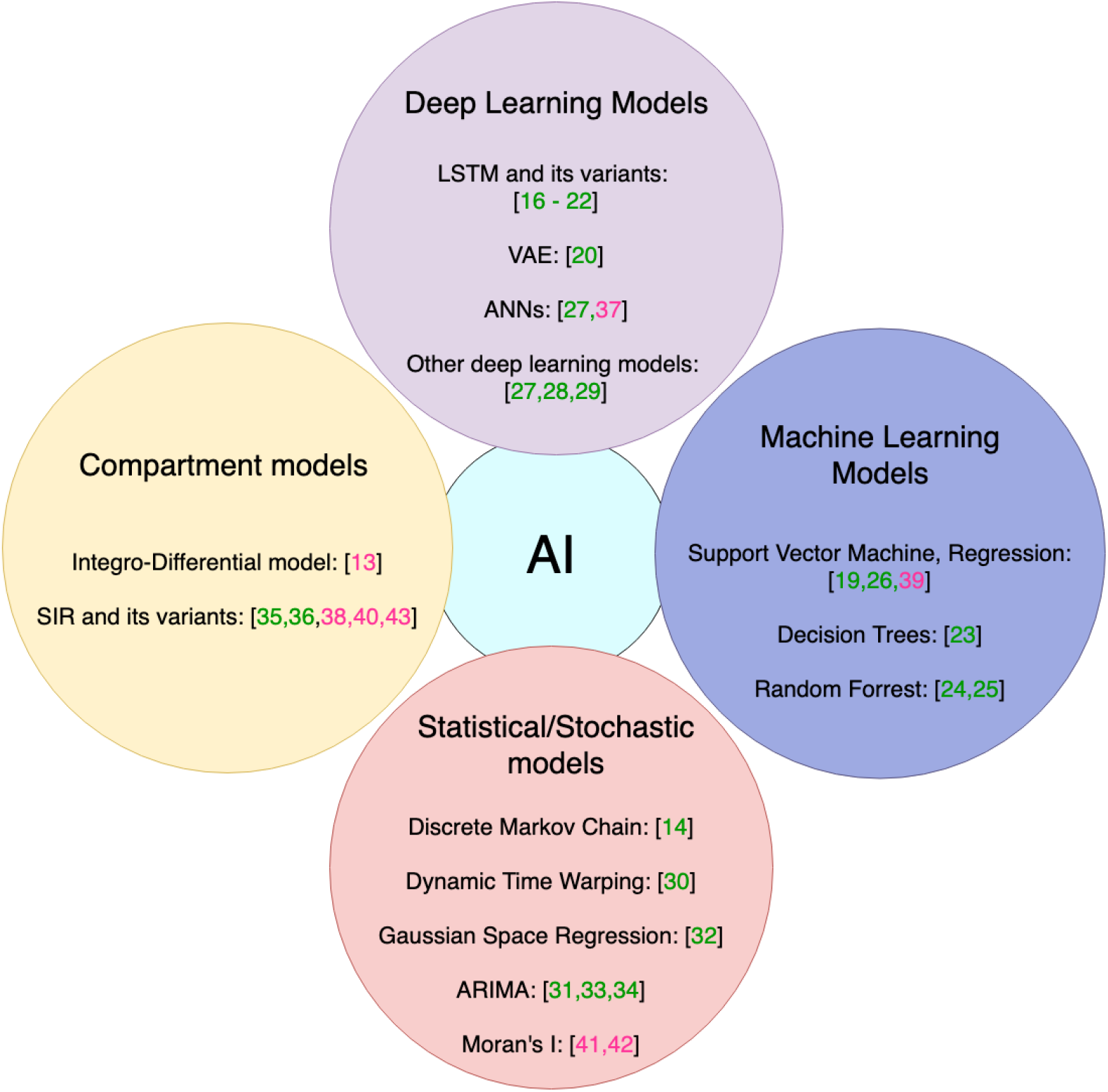
AI models used in spatio-temporal modeling and forecasting. The reference number for each model is given in square brackets. Green- temporal model, Pink- spatio-temporal model.

#### Temporal modeling

Temporal forecasting of COVID-19 infection cases have been evaluated using multiple deep learning models. A multi-step prediction using the neural network models - Long Short Term Memory (LSTM), Convolutional LSTM (ConvLSTM), Bidirectional LSTM (Bi-LSTM)- as a case study for Canada,^16^ different states in India,^17^ and USA^18^ have been studied. The Bi-LSTM model and the ConvLSTM models are shown to outperform the other LSTM variants with a Mean Absolute Percentage Error (MAPE) of 3% for day ahead predictions. Accuracy of variants of LSTMs are further compared with Gated Recurrent Units (GRU), a Variational Auto-Encoder (VAE), Convolutional neural networks (CNNs), and Support Vector Regression (SVR) models.^19, 20, 21^ The models are evaluated on multiple countries where substantial data is available (USA, Italy, Spain, China, Australia) to validate that the Bi-LSTM and VAE (MAPE less than 3%) models outperform the conventional models. Meanwhile, a data driven forecasting method has been developed to estimate the number of positive cases of COVID-19 in India for the next 30 days.^22^ The number of recovered cases, daily positive cases, and deceased have also been estimated using LSTMs and curve fitting. In order to handle the data imbalance problem, a decision tree algorithm is developed to extract rules to determine the optimal algorithm for forecasting.^23^ In addition, machine learning based random forest model,^24, 25^ Support Vector Machine(SVM) model,^26^ Artificial Neural Networks (ANNs)^27^ and other deep learning models^28, 29^ have been used in forecasting.

A novel statistical method has been developed to forecast the spread of COVID-19 globally. This method considers lead-lag effects between different time series using the dynamic time warping technique to analyze non linear relationships among nations.^30^ Thus, it was able to determine underlying causal relationships such as the origin of the virus and the forerunner in given regions. A Non linear Autoregressive Artificial Neural Networks (NAR-ANN) and Auto Regressive Integrated Moving Average (ARIMA) model was used to forecast the spread of COVID-19 in Egypt.^31^ Furthermore, an analysis of COVID-19 deaths using reduced space Gaussian process regression has shown a correlation coefficient of 98·9%.^32^ In an attempt to solve the patient triage problem related to COVID-19, researchers in Italy have formulated a discrete time markov chain model to predict the number of COVID-19 cases, to thereby, efficiently allocate ICU resources across the country.^14^ The critical need for COVID-19 forecasting comes from the need to understand how to implement containment strategies while balancing its impact on the country’s economy and thereby the livelihood of people. From an economical impact standpoint, a modified ARIMA model was developed to forecast the number of COVID-19 cases and the stock market in Spain^33^ for the given period. ARIMA has also been ensembled with Wavelet Transforms to model stationary and non-stationary trends using a 10-day forecast.^34^

Epidemic modeling in the past has extensively used compartment models, especially the Susceptible - Infective - Removed (SIR) model and its variants. Researchers in China have implemented a dynamic Susceptible Exposed Infectious Removed (SEIR) model along with an Artificial Intelligence model to predict the COVID-19 spread in China and identify underlying patterns in the spread.^35^ A modified SEIR model has been implemented to forecast the spread of COVID-19 and its burden on hospital care under varying social distancing conditions.^36^ This study concludes that the best parameter to assess the effectiveness of confinement and risk of virus diffusion, is the average number of daily contacts in a population (*c*).

#### Spatio-temporal modeling

Researchers have focused on modeling and forecasting the spread of COVID-19 in the spatio-temporal domain to identify not only ‘‘how much” the disease has spread but also ‘‘where” it has spread to, and to ‘‘what extent”.^37, 38, 39^ A study conducted for Germany has developed a memory based integro-differential network model to predict spatio-temporal outbreak dynamics such as number of infections, hospitalization rates, and demands on ICU due to COVID-19 at state level.^13, 40^ The model takes into account effects of different containment strategies, event and contact restrictions, and different courses the infection may take; which is not possible when dealing with traditional SIR models. A novel approach is taken using Moran’s I test to find regional correlations of COVID-19 cases in China.^41, 42^ The spatial association between states are modelled into the following six types:^41^ 1) Sharing of borders 2) Euclidean (Shortest) distance 3) Population 4) Population density 5) Number of doctors and hospitals 6) Number of medical beds. Results show high positive correlation between an infected region and its adjacent regions for the first five models. This method is useful for predicting outbreaks in specific regions, which in turn will allow policymakers to take proactive measures to prevent COVID-19 spread to adjacent regions. The county level spread of COVID-19 in relation to the healthcare capacities in Ohio, USA is predicted and the results show that the disease spreads much faster in counties that facilitate air transportation to other counties.^43^ This shows that similar to attributes such as population, underlying spatio-temporal attributes such as inter-state travel can be employed to predict the dynamic spread of the pandemic.

### 3.2. Modeling the impact on diverse populations

In this section, we review methods that group the population into clusters based on different factors such as occupation, age and other such parameters and analyze COVID-19 spread in those demographics. An overview of what characterizes the population diversity is given in Fig. 4.

**Figure 4:**
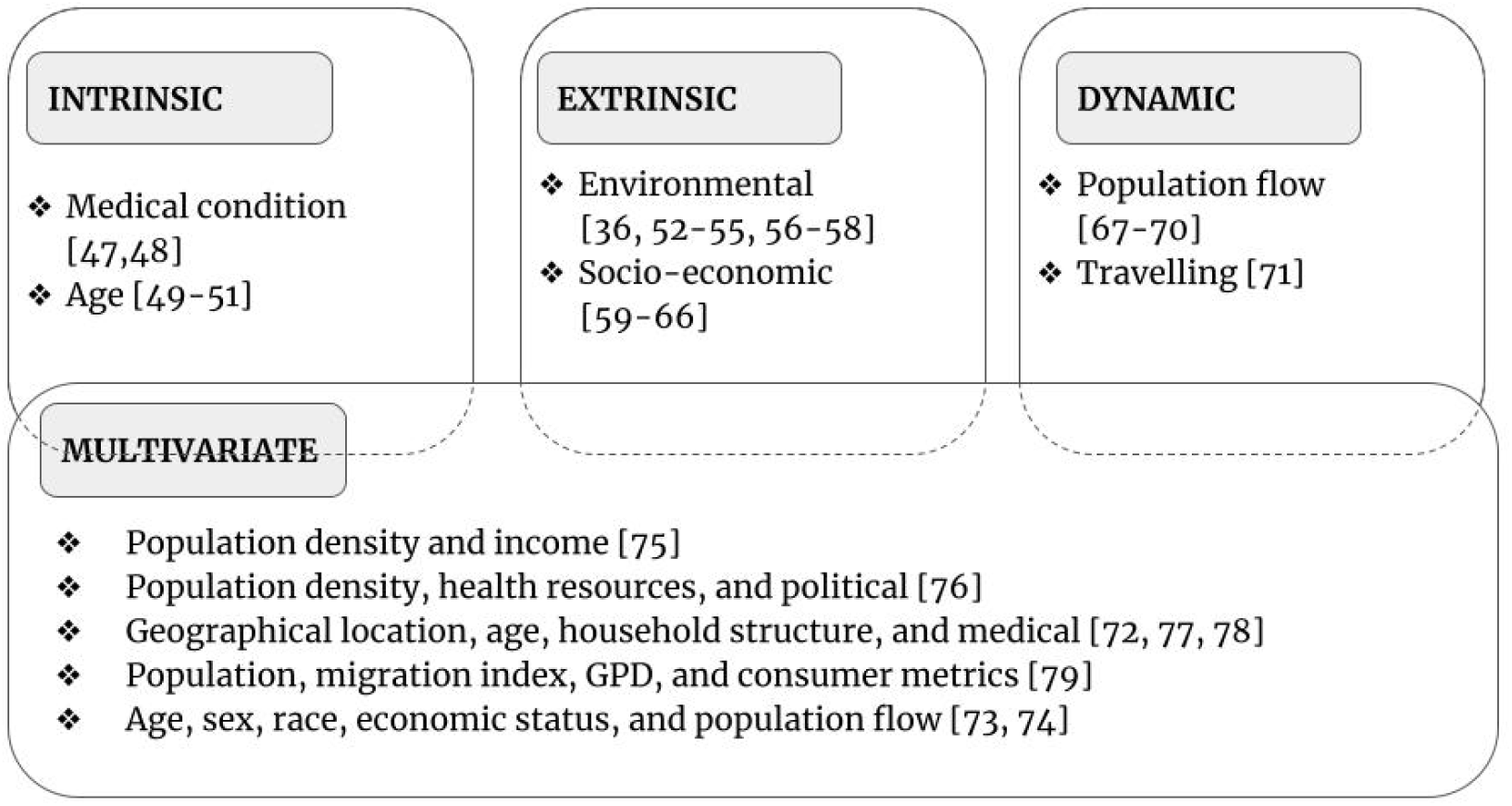
Impact Modeling. The reference numbers of research papers are given in square brackets.

The importance of this diversity/context awareness of information for handling the pandemic across the globe has been studied^44, 45^. Even within similar geographies, socio-economic factors have been shown to have an effect on the spread and severity of the pandemic.^46^ This work shows why the analysis, prediction and mitigation of COVID-19 related issues should be done considering the context (diversity of the population) for best results.

Models developed to handle diversity awareness are mostly mappings between higher dimensional spaces (in cases where the context is passed as an input) with the capability to capture the multi-modal behavioral of data. We see multi-variate statistics, time series models, artificial neural networks and other AI models being used in this landscape. Key challenges observed in this domain are, handling non-uniformly sampled data, incomplete data and figuring out the context without it being readily fed into the models.

The parameters taken into account to characterize diversity are analyzed in the following subsections (as given in Table 1). A summary of this section is given in Table 2.

**Table 2:**
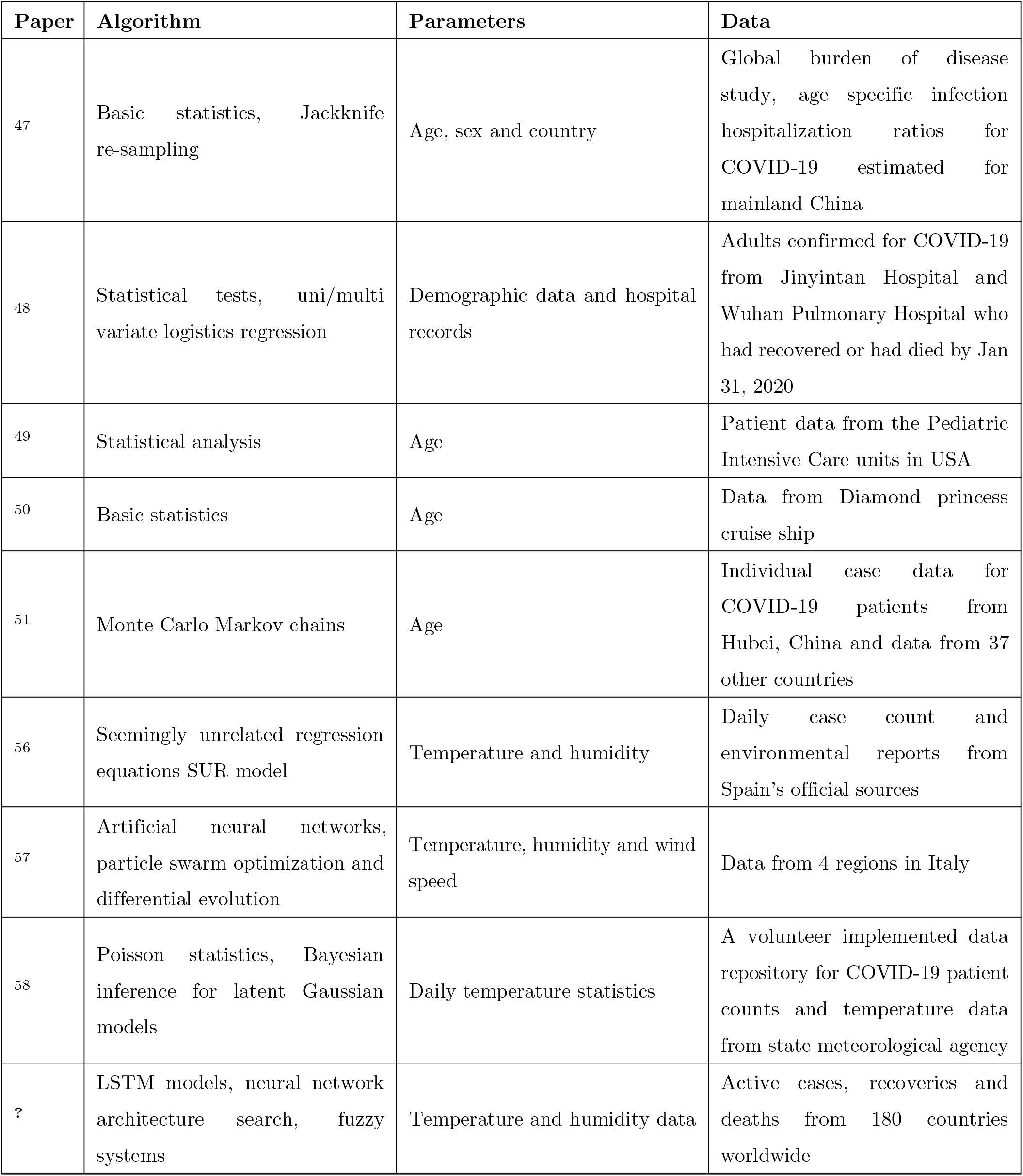
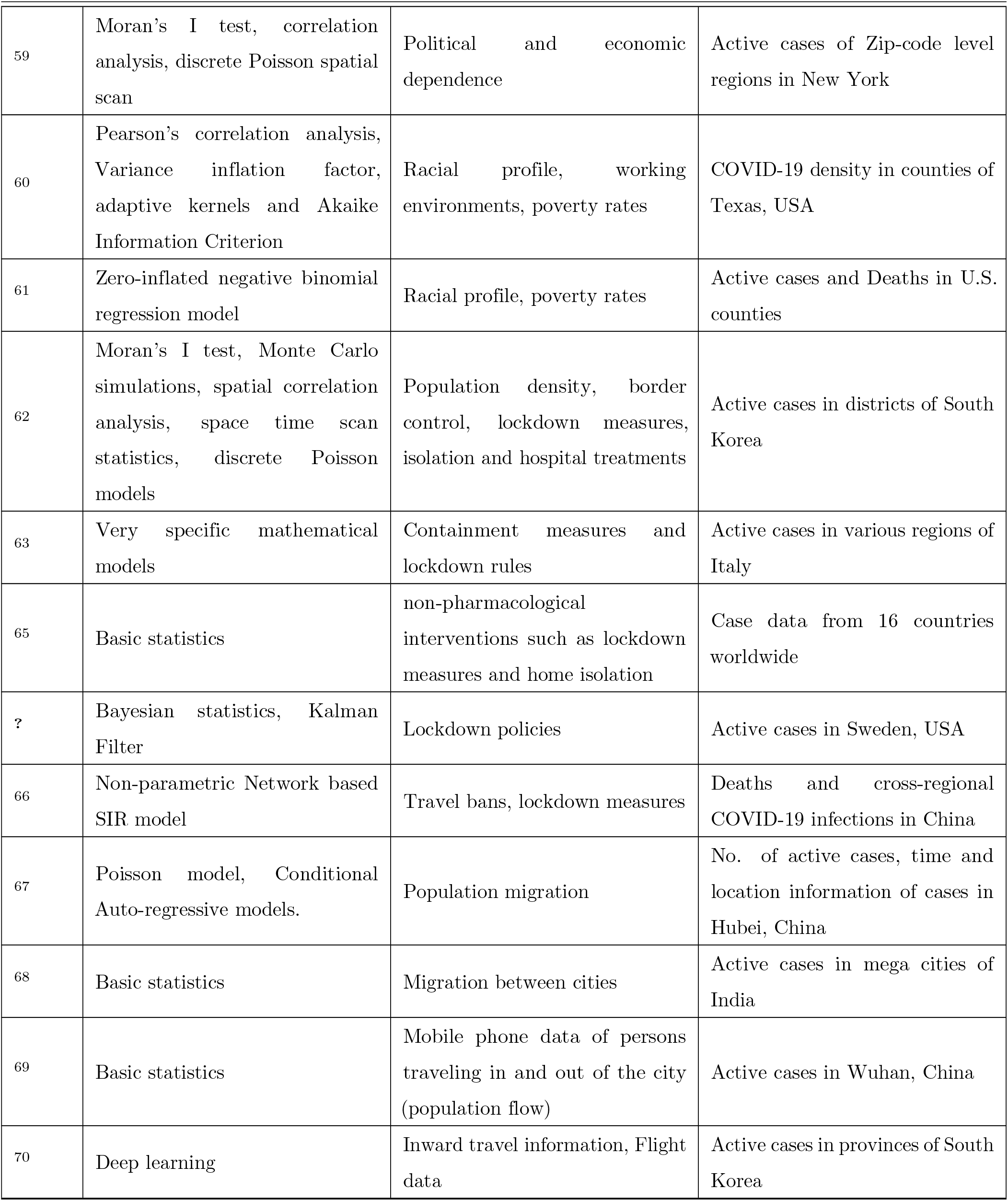
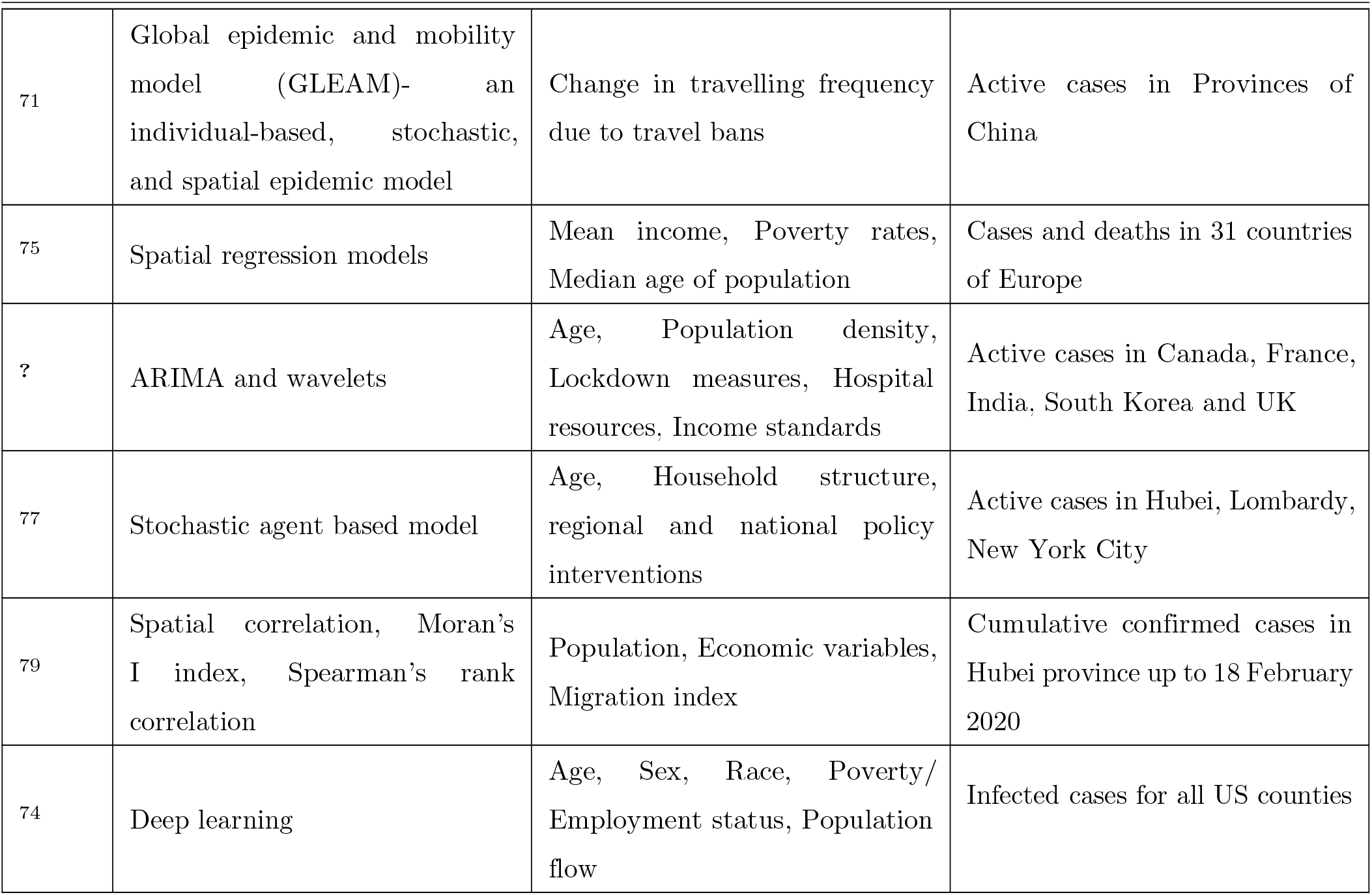
Summary: Diversity aware AI

#### 3.2.1. Intrinsic Parameters

##### Medical Conditions

A large-scale analysis of the severity of COVID-19 among the population, given the existing statistics of the medical conditions as per the Global Burden of Disease and Population census among the world, is done using statistical analysis coupled with resampling techniques.^47^ This study shows the importance of utilizing larger data sources to generate insights during the pandemic. Another study aiming to uncover a similar correlation between pre-existing conditions and COVID-19 mortality was conducted on individual patient data from Wuhan using linear regression and statistical techniques.^48^

##### Age

In this section, we discuss the research work attempting to uncover the differential impact of COVID-19 on different age groups. The age dependence of COVID-19’s impact on children is studied using statistical analysis to thereby generate predictions on the population as a whole by considering data from USA.^49^ This work goes on to generate recommendations based on the predictions on how best to prepare for large scale outbreaks. The age stratified CFR rate (case fatality rate -- the ratio of deaths to the number of total cases) is analyzed using basic statistics.^50^ Another study performs an in-depth analysis on a larger dataset from around the world (China’s statistics coupled with the rest of the world).^51^ The study also sheds light into the age dependence of healthcare in an overwhelmed system. From the literature, it is concluded that age dependence is mostly analyzed using classical algorithms, as opposed to more modern techniques.

#### 3.2.2. Extrinsic Parameters

##### Environmental differences

The spread of COVID-19 has been analyzed with regard to environmental factors such as temperature, precipitation and humidity.^52, 53, 54, 55^ One study uses a spatial Seemingly Unrelated Regressions (SUR) to model the COVID-19 spread as an inter-regional contagion process.^56^ This does not show the effect of temperature and sunshine on the behavior of the virus, but instead analyzes the people’s behavioral changes based on environmental factors, and thereby the spread of the virus as a result of these changes. Another similar study (using ANN based differential equations and particle swarm algorithms) shows that population density (followed by relative humidity) has a higher impact on the spread of COVID-19 in comparison to temperature and wind speed.^57^ Statistical models have also been used to show the absence of a correlation between the daily temperature and case counts in Spain.^58^ Similarly, the performance of ANN based COVID-19 prediction is evaluated to show how absence of weather data does not make a difference.^29^

##### Social, Economic and Political factors

Moran’s I test is widely used to analyze spatial correlations in pandemics including social, economic and political dependencies. One such study computes the correlation between testing and COVID-19 positive rates, and the socio-economic status in zip code level clusters of New York. The study recommends healthcare resources to be directed towards areas where low testing and high positive rates are prevalent.^59^

Another study explores spatial relationships between socio-demographics and the spread of COVID-19 in three regions of Texas using ordinary least squares (OLS) and geographically weighted regression (GWR). The authors quantify the influence of poverty on COVID-19 infection density for a given region. In turn, it helps policy-makers direct healthcare resources and provide more emphasis towards low-income regions in a state.^60^ The effect of the percentage of African-American population on COVID-19 disease and death rates, for a given county in the United States is analyzed. Results indicated that nearly 90% of counties with a disproportionately higher number of African-Americans had a higher percentage of cases and deaths. The relationship between this socio-demographic and COVID-19 was fit to a Negative Binomial Regression (NBR) model, which can forecast the number of future cases in relation to income and racial parameters in a given county.^61^

A case study of South Korea evaluates lockdown policies to find noteworthy COVID-19 clusters in space and time. It was observed that clusters were contained faster as time progressed (i.e. earlier outbreaks took time to contain but clusters that started later were contained quickly). This was evidence of how government policies can affect the spread of COVID-19. The model also took into account the population density of districts (higher population density implies higher disease spread).^62^ Similarly, time, space and population demographics have been used to model the COVID-19 outbreak containment in Italy.^63^ A prediction modeling study to measure the effect of Non-Pharmaceutical-Interventions (NPI) on the spread of COVID-19 has been conducted for USA at the state level^64^ and for 16 countries from diverse geographical regions.^65^ The authors analyze the effects of no intervention, cyclic mitigation or suppression measures followed by relaxation period. The effects on ICU admissions and death is evaluated for these different measures. A similar study has been conducted for North America, Sweden^15^ and China,^66^ as they have taken contrasting policies to battle COVID-19.

#### 3.2.3. Dynamic Parameters

##### Travelling and Population flow

Focusing on travel and mass population flow, a recent study found correlations between the number of persons migrating from Wuhan to other cities and the number of cases in those cities. Using this, a risk analysis was developed using a Bayesian space-time model to quantify the risk of a person migrating from Wuhan spreading COVID-19 to another population.^67^ While the similar scenario in India’s mass migration has been studied,^68^ AI is yet to be used in the Indian situation. Dynamic Parameters have also been used to analyze the spread of COVID-19.^69^ This study identifies the regions with high risk of early transmission, and forecasts the distribution of COVID-19 cases from Wuhan, using statistical methods on population flow data.

Meanwhile, modeling the effect of population flow with consideration on to the geography has outperformed the naive population flow models in terms of accuracy. The former was done by a neural network that can absorb time series information of the patient counts and travel statistics, as well as knowledge about the geographical hierarchy of countries and continents.^70^ A study further discusses the impact of travel restrictions on the spread of COVID-19 on a regionally connected population. To identify the spread of the disease, a Bayesian approach is used. The results show that travel restrictions had a minimal effect unless they were supplemented with other behavioral changes to mitigate the disease spread.^71^

#### 3.2.4. Multivariate inputs

The methods in these articles attempt to model the complex relationships between different parameters and uncover correlations that might not be obvious in traditional statistical analysis.^72, 73^ These models are designed based on both human intuition and AI. Understanding the inter-dependence of the input variables used to model the spread of COVID-19 is important in developing an intuitive deep learning model. Combining both stationary and time dependent parameters, a deep learning architecture “CovidNet” has been proposed to take advantage of the different types of inputs. The work also shows the importance of understanding the inter-dependence of the input variables to develop deep learning models coupled with human intuition.^74^

To identify the socio-economic variables most influential towards COVID-19 fatalities, a spatial regression approach has been used.^75^ Out of 28 possible demographics; population density, income, and poverty rates were found to have the highest correlation with infection rates and fatalities in a given region. This study also models fatalities of a given region using these reduced socio-economic variables. The reduction of variables vastly helps real-time forecasting due to simplicity and reduced computational cost. Another study developed a real-time risk assessment method for future cases in terms of fatality rate of an affected country, using population density, healthcare resources, and government enforced preventive measures. The number of future cases was predicted using an ARIMA model, and a regression tree algorithm was used to validate the argument that the aforementioned variables have the highest impact on fatality rates in those countries.^76^

Modified SEIR models including time-dependent parameters have been explored analysing multiple categories such as age, sex, household structure, current medical conditions and spatial locations.^77, 78^ The study on individual level model for COVID-19 transmission spread in the city of Lombardy suggests that salutary sheltering of persons belonging to different age groups along with physical distancing measures may significantly reduce the rate of COVID-19 transmission. A study of COVID-19 spread in Hubei, China analyzes the spatio-temporal spread of COVID-19 using spatial autocorrelation and Spearman’s rank correlation methods. From a number of demographics; resident population, migration index, Gross Domestic Production (GDP) and Total Retail Sales (TRS) of consumer goods were determined to have a strong influence on the spread of COVID-19.^79^

## 4. Discussion and potentials of AI for future avenues

The literature survey has uncovered two key application areas in which AI is used-- predicting the future impact of COVID-19 on populations and uncovering the differential impact of COVID-19 on diverse segments of the population. The performance of predictive algorithms have been limited by a range of factors (Section 3.1). Impact modeling has generated interesting insights on the virus as well as the society at large (Section 3.2).

Despite the recent growth of AI, many existing solutions rely on traditional methodologies, without any automation. Even though medical systems have been computerized in the past decades, the unprecedented nature of COVID-19 has brought forward issues that need to be automated but are still dependent on human intervention. For example, patient care and scheduling medical personnel are mostly done through a manual rule-based methodology to ensure that the people that are more infectious are handled with rigorous spread prevention measures.^80^ The application of AI for such novel issues poses certain challenges, but overcoming these challenges and exploiting the available methodologies could result in a marked improvement in a variety of fields, especially within the medical domain. Table 3 lists a few COVID-19 related issues for which AI may be an ideal candidate.

**Table 3:** Potential of AI

| Problem | Research gap | AI methods |
| --- | --- | --- |
| Identify reasons for high mortality rate caused by COVID-19 . | Traditional techniques based on simple statistical models cannot identify complex clusters. | Deep clustering based on convolutional networks can be used for complex clustering. |
| Find the correlation between COVID-19 and socio-economic problems. | Classical methods do not consider complex feature sets. | Deep clustering can be used as an alternative to identify complex features. |
| Contact tracing through mobile applications. | A major drawback is their inability to utilize Big Data efficiently. | The increased processing power of mobile devices enables the use of AI to improve efficiency and performance. |
| Forecast COVID-19 cases and deaths based on spatio-temporal data. | Certain models fail to consider spatial and temporal relationship of data. | Temporal information can be modelled effectively through Graph Neural Network and transformers. |
| Find correlation between disease spread and population behavior such as travelling patterns. | Classical techniques use individualistic data and do not consider the related features between individuals. | Spatial information can be modelled using network models such as *graph neural network* to identify social clusters. |
| Social distancing based on computer vision. | Environment-specific risk assessment is an issue which has not been studied in detail. | Existing CNN based solutions can be aggregated and extended for risk assessment. |

One major drawback of traditional statistical techniques is their inability to take advantage of Big Data. For example, statistical techniques have often been used to find the most influential factors that leads to COVID-19 related deaths,^48^ and the socio-economic issues that arise as a result of COVID-19.^81^ While statistical studies are helpful, identifying specific clusters of features will be instrumental for decision making. Deep clustering techniques based on convolution neural networks have outperformed traditional methods^82^ for such clustering problems. In addition to handling complex input data, the convolutional models learn spatial information and correlation that results in better performance.

Similarly, traditional techniques do not handle Big Data efficiently. For example, contact tracing through the use of mobile application such as ‘‘WeChat’’ has been effective in many countries.^83^ However, one major limitation is their inability to ingest a large volume of data for decision making and adapt accordingly.^84^ Due to the improvement in processing power of mobile devices, AI algorithms can be adapted to solving problems such as proximity identification and improve the accuracy and reliability of these systems through adaptation and real-time learning.

Furthermore, classical techniques often fail to model the spatio-temporal relationships of the input data. Studies have been conducted to find the correlation between the traveling patterns of individuals and how it correlates to the spread of disease.^83^ However, individual-level tracking only focuses on geographical location and overlooks other factors such as social clusters, race and age. It would be interesting to consider these social clustering parameters for this study, as they may have significant correlation with the spread of pandemic. Graph Neural Networks (GNN) can be used to model and identify specific events in the spatial and/or temporal domain. The spatial information can be modelled through graphs and the information can be propagated within the nodes through message passing. In addition to GNNs, using state-of-the-art NLP techniques like Transformers,^85^ spatio-temporal information that pays specific attention to events (e.g. sudden spikes in patients) in the temporal and/or spatial domain can be efficiently modeled.

Despite the large number of recent vision-based solutions for COVID-19,^86^ the risk level assessment for a particular environment is neither defined nor addressed properly. Comparing different environments (using a combination of vision techniques for social distancing, face mask identification and other visible metrics) and quantifying the risk of transmission in those particular environments would be helpful in making decision such as implementing social distancing measures and easing/strengthening problem.

## 5. Limitations and Conclusion

The rapid development and growing research in AI is amassed as preprints rather than peer reviewed work. Thus, the exclusion of preprints limits the scope of the review process. In addition to the exclusion criteria, the setting of AI research in the contemporary world imposes restrictions to review. A large portion of annual investment (US$ 1·7 billion in 2018^87^) for R&D in the intersection of AI and healthcare comes from tech corporations such as Google and Facebook.^88^ Since a major portion of the AI research and development ends up in commercial deployment, they are not made available to the public.

It is important to note that our recommendations in Section 4 were built on the premise that the methods we prescribed were not already explored by other researchers. Perhaps, the methods were implemented and not published because they did not yield the expected positive results.

Moreover, our review only focuses on papers written in English whilst the outbreak of COVID-19 started from China. There are many research articles and reports written in Chinese languages that are not considered in this review. Furthermore, India recorded a large number of COVID-19 patients as well and it’s demographics and socio-economics presented a wide range of unique challenges (mass immigration, social inequalities in access to healthcare) in handling the situation. Analysis and reports on this matter that were done in local languages are also not considered in this review.

AI has been deployed towards the combat of COVID-19 all around the globe. The severe acute respiratory syndrome and its novel nature of spread has required an effective pandemic response. AI-driven modeling and forecasting systems have been successful in alleviating the situation.

In this review, we have explored the gaps of AI usage as a tool to fight against COVID-19. Specifically, (a) existing methodologies in the pandemic response could be replaced by new AI technology in modeling and forecasting, (b) new AI technology could be introduced to surpass prevailing AI modeling and forecasting technique outcomes, and (c) new AI techniques in various other fields that have not been experimented in a pandemic situation before could be drawn parallel to the COVID-19 response in an effective manner.

## Data Availability

The data used in this study are openly available on the internet.

## Author Contribution

GJ, JH, UM, RP, SS, HW, ME, RG, PE, VH, DG, AR, SD, and JE conceptualized the study and designed the methodology. RG, PE, VH, DG, AR, SD, and JE supervised the project. GJ, JH, UM, RP, SS, HW, and ME carried out the literature search, reviewed the articles, and wrote the draft. RG, PE, VH, DG, AR, SD, and JE reviewed and edited the draft. GJ, JH, UM, RP, SS, HW, ME, RG, PE, VH, DG, AR, SD, and JE had access to the data, read the final manuscript and approved it.

